# A Novel Convolutional Neural Network for COVID-19 detection and classification using Chest X-Ray images

**DOI:** 10.1101/2021.08.11.21261946

**Authors:** Muhammad Talha Nafees, Irshadullah, Muhammad Rizwan, Maazullah, Muhammad Irfanullah Khan, Muhammad Farhan

## Abstract

The early and rapid diagnosis of severe acute respiratory syndrome coronavirus 2 (SARS CoV-2), the main cause of fatal pandemic coronavirus disease 2019 (COVID-19), with the analysis of patients chest X-ray (CXR) images has life-saving importance for both patients and medical professionals. In this research a very simple novel and robust deep-learning convolutional neural network (CNN) model with less number of trainable-parameters is proposed to assist the radiologists and physicians in the early detection of COVID-19 patients. It also helps to classify patients into COVID-19, pneumonia and normal on the bases of analysis of augmented X-ray images. This augmented dataset contains 4803 COVID-19 from 686 publicly available chest X-ray images along with 5000 normal and 5000 pneumonia samples. These images are divided into 80% training and 20 % validation. The proposed CNN model is trained on training dataset and then tested on validation dataset. This model has a promising performance with a mean accuracy of 92.29%, precision of 99.96%, Specificity of 99.85% along with Sensitivity value of 85.92%. The result can further be improved if more data of expert radiologist is publically available.

## 1 Introduction

The first case of novel coronavirus (COVID-19) reported in December 2019 at Wuhan city in China [1, 2]. This infectious disease caused by the SARS CoV-2 having high degree of spreadability which is a great challenge for economic and healthcare system. It also disturbed the education sector, the social life; in short every walk of life in a very devastating way. Recently in January 27, 2021 this fatal disease has more than 100,922,270 cases and 2,169,466 confirmed deaths [3]. To diagnose it on time and then taking necessary actions accordingly, is essential to control this fatal viral pandemic. The standard method for the detection of COVID-19 listed by WHO is Reverse transcription-polymerase chain reaction (RT-PCR) which detects the presence of the virus but later it is reported that RT-PCR has variable/low sensitivity [4]. In addition, it requires a lot of time and need much expensive testing kits and expert medical professionals which are not easily available at large scale. Therefore, researchers and experts are focusing on an alternate ways for the early detection of COVID-19. As the common clinical feature of severe COVID-19 infection is ground glass opacity and consolidation [5-7] therefore radiological images of chest X-rays and CT images is a useful diagnostic tool for assessing COVID-19 disease [8-10] but interpretation of the images is challenging and time consuming [11]. Fortunately, the deep learning models overcome these challenges by images recognition and classifications [12, 13]. The prominent Convolutional Neural Networks (CNN) models such as Alexnet, VG-GNet, ResNet and Inception perform well in practical image recognition and classification [14-16]. It also shows satisfactory performance in solving the medical image classification [17-19]. Therefore, it plays a vital role in the early detection of COVID-19 disease and save both time and money. Currently the available dataset are either CT images or chest X-Rays but CT images are expensive and more sensitive to pulmonary disease. However, CXR is readily available in every hospital at very low cost. In this research a simple, novel and robust deep-learning convolutional neural network (CNN) model is proposed for COVID-19 classification with publicly available chest X-rays images. Section II describes the related works. Section III describes the dataset. The proposed model and result are described in Section IV and V respectively and the concluding remarks are given in Section VI.

## 2 Related Works

Extensive research work is going on for determining the accurate and reliable deep-learning (DL) model for the detection and classification of COVID-19 disease. Mostly researchers classify chest X-ray and CT images of patients by using different deep-learning techniques. Apostolopoulos and Mpesiana proposed ConvNet model for COVID-19 detection with two different publicly available X-ray images. In this research most frequently pre-trained networks are used i.e. VGG19, MobileNet V2, Inception, Xception and Inception ResNet V2 and evaluate on the basis of several performance metrics such as accuracy etc. and concludes that MobileNet-V2 and VGG19 perform well than other pre-trained ConvNets [20]. Similarly, Narin et. Al proposed three pre-trained network that are Inception-ResNetV2, InceptionV3 and ResNet50, for COVID-19 patients with CXR and achieved the accuracy of 87%, 97% and 98% respectively [21]. These models are only trained on 50 COVID-19 and 50 normal CXR therefore the accuracy might be declined for large training data. Zhang et al. proposed a deep learning network for detecting COVID-19 patients trained on 100 COVID CXR while the model is pre-trained on different domain dataset (ImageNet) having accuracy of 96% and 70.65% for COVID and Non-COVID respectively [22]. Sethy and Behea also proposed DL model based on ResNet50 plus SVM for Coronavirus detection [23] which achieved the accuracy and F1-score of 95.38% and 91.41% respectively. In another research [24] a small set of X-rays images are used to tune a pre-trained ResNet50 and VGG 16 with overall accuracy of 91.24%. Li et al. proposed a model architecture named COVNet with sensitivity, specificity and AUC of 90%, 96% and 0.96 respectively [25]. Wang and Wong proposed a COVID-Net model to classify Normal, pneumonia and COVID with accuracy of 92.4%[26]. Similarly, other researchers also put efforts to detect COVID cases from radiographic images using different DL techniques in [27-30]. Most of the research work discussed regarding COVID-19 detection use transfer learning which use pre-trained base models as a part of their model architecture, which is usually pre-trained on ImageNet dataset, which is not a medical domain dataset and the result might not be reliable. The remaining models use very small number of training radiological images which cause overfitting. Furthermore those datasets are mostly imbalanced. All the said causes may decline the performance of both binary and multiclass classifier. Hence, In this research a custom CNN model having simple architecture and less number of trainable parameters is proposed for COVID-19 classification. This model is trained from the scratch with balanced augmented dataset.

## 3 DataSet

### 3.1 DataSet

The dataset consist of a total 10,686 images. These images are collected from seven different publicly available databases that are Kaggle COVID-19 chest X-ray image Dataset[31], Kaggle RSNA Pneumonia Detection Challenge Dataset[32], SIRM COVID-19 Database[33], GitHub COVID-19 Image DataCollection[34], GitHub Covid Chest X-ray Dataset[35], Kaggle Covid-19 Chest Radiography Database [36], and GitHub Actualmed Covid Chest X-ray Dataset[37]. The 10,686 images consist of three classes i.e. normal, pneumonia and COVID-19. Table 1 explains the distribution of Images among these three classes. In each class the images are further divided into Train for training purpose and valid for validation purpose with 4:1. The distribution of data shown in Figure 1 indicates that the publicly available X-ray images for COVID are less and also imbalanced. To overcome this problem several augmentation techniques are used.

**Table 1:** The distribution of Images among three classes.

| Class | Train | Valid | Total |
| --- | --- | --- | --- |
| COVID | 549 | 137 | 686 |
| PNEUMONIA | 4,000 | 1,000 | 5,000 |
| NORMAL | 4,000 | 1,000 | 5,000 |
| Total | 8,549 | 2,137 | 10,686 |

**Fig. 1:**
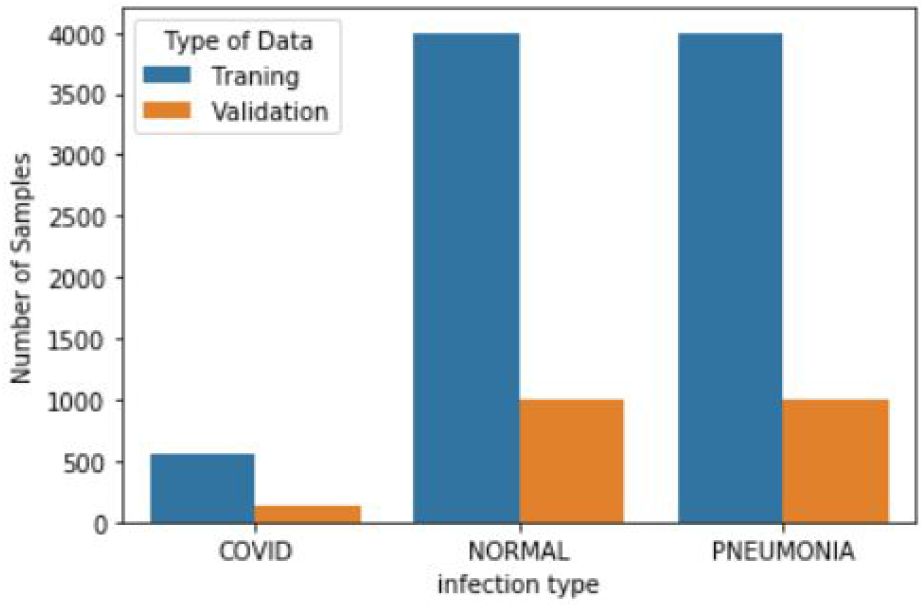
Distribution of images before augmentation.

### 3.2 Preprocessing

Before feeding data to CNN model, the images are normalized to prevent the model from saturation and to enhance the training process. In addition, the images are resized from 1024x1024 to 224x224 in order to decrease the computational cost. Furthermore, the images are shuffled to make the data more versatile which eventually leads to generic training and the model will become more generalized. Several augmentation techniques are used to increase the COVID X-ray images and to make the dataset balanced. These techniques are Top/Centre cropping, salt noise, horizontal and vertical flipping, rotation of 15o and brightness control as shown in Figure 2.

**Fig. 2:**
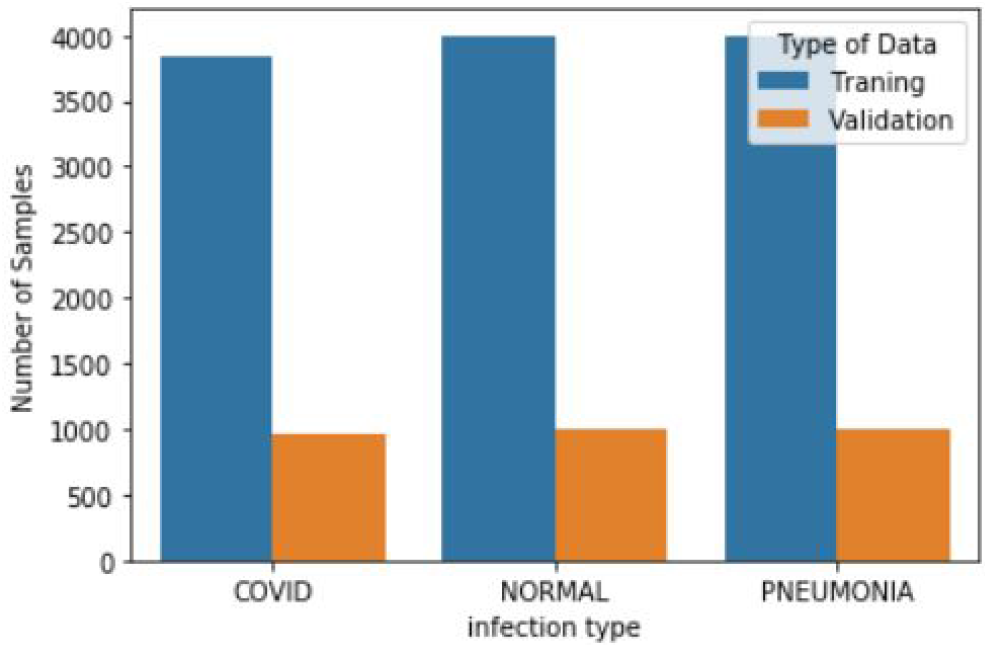
Distribution of augmented images.

These augmentation techniques are useful to avoid under-fitting because the system is recognizing each image as a new entity. Furthermore, the dataset become balanced and the number of COVID-19 train images increased from 549 to 3844 samples and validation samples increased from 137 to 959 as shown in Figure 3.

**Fig. 3:**
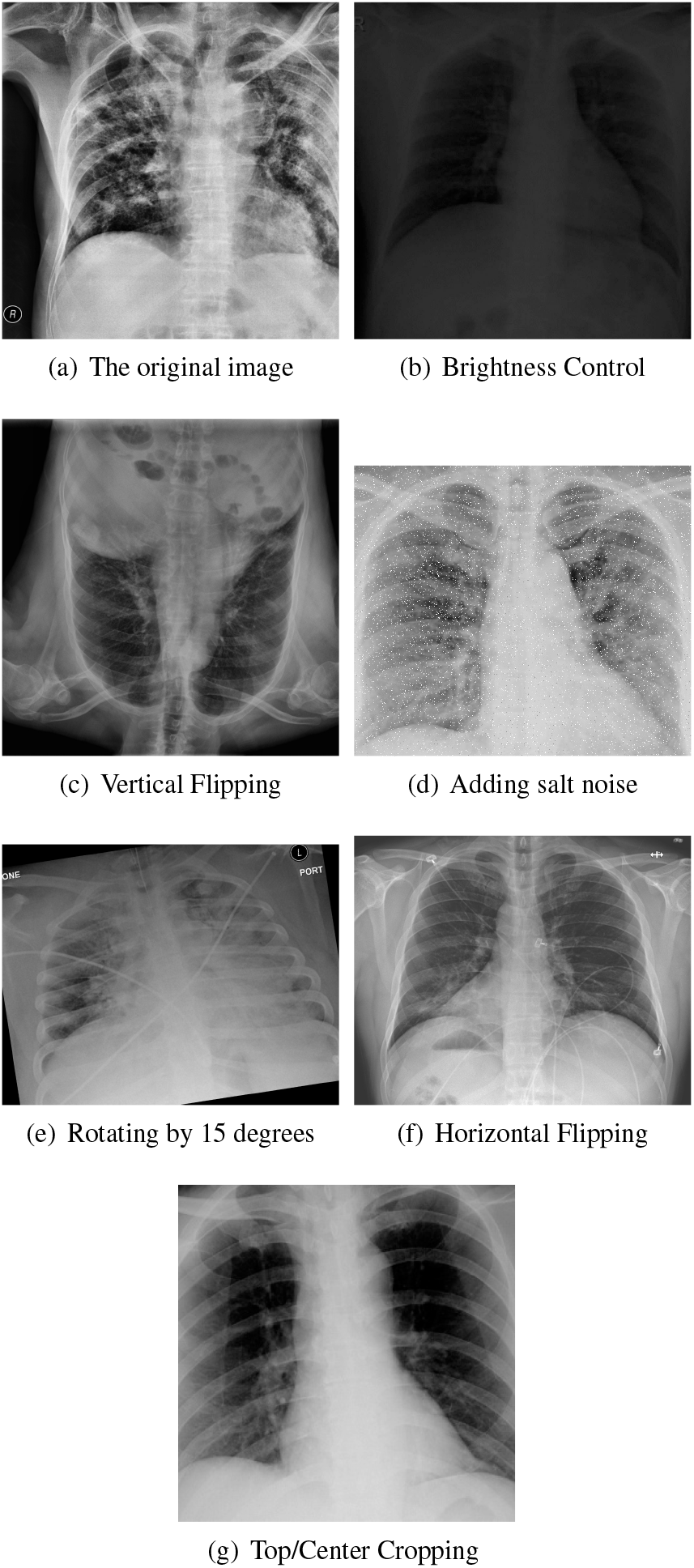
Augmentation techniques

### 3.3 Proposed Model

The proposed CNN network is comprised of 16 layers having three convolutional layers in which each convolutional layer is followed by max-pooling layer while a dropout layer is used after second and third max-pooling layer. After the dropout layer is the flatten layer which is followed by two dense layers in which the first one is followed by dropout layer which is further followed by batch normalization layer and at the end softmax layer is used for classification. Furthermore, in each convolution layer and in first dense layer Relu activation function is used as shown in Figure 4.

**Fig. 4:**
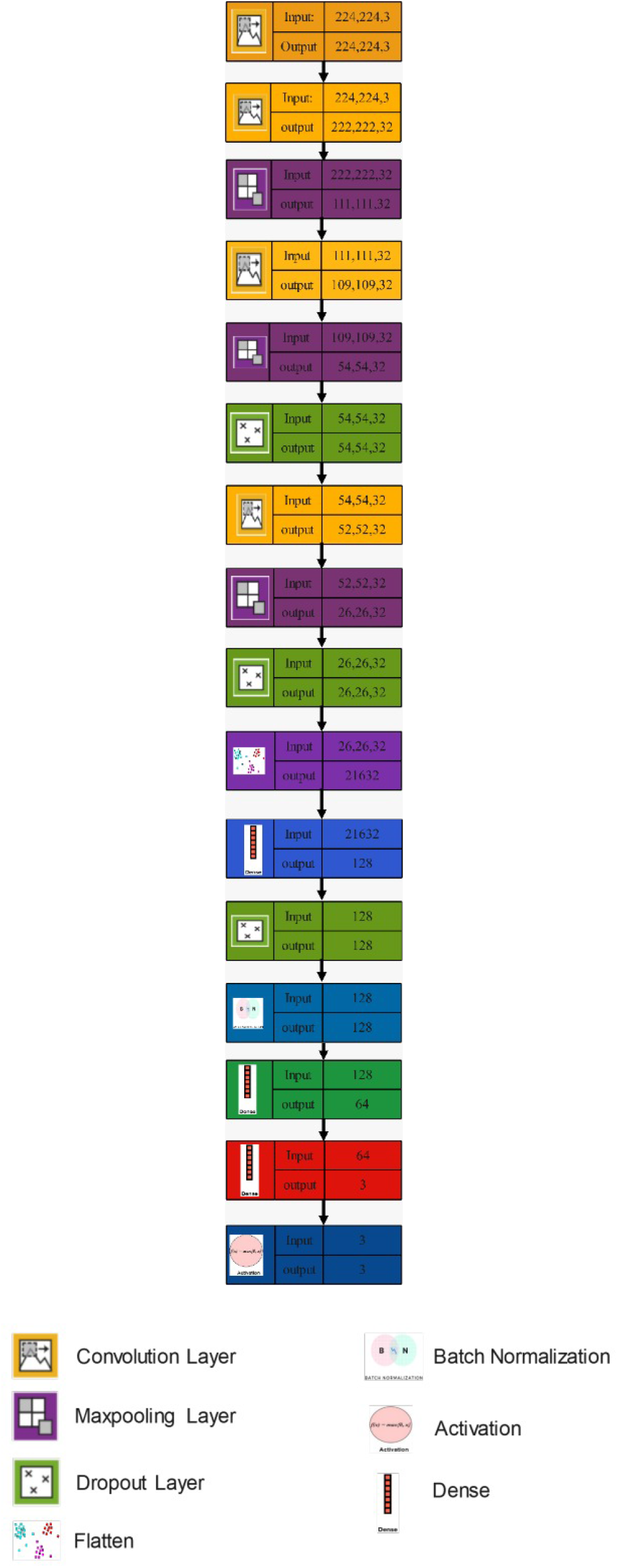
Architecture of proposed customized CNN model.

Each convolutional layer has 32 filters of size of (3 x 3) for features extractions while in max-pooling layer a (2x2) window size is used. It is used to reduce the size of the features map for decreasing the computation. The three dropout layers are used with dropout of 0.5 to reduce the overfitting by randomly dropping some nodes in each step. The feature extracted by convolutional network are flattened into 1-dimensional vector so that the fully connected network classify them using softmax into three classes i.e. COVID, Normal and Pneumonia while the batch normalization layer is used to standardize the input of each batch which increases the speed of training. The detailed parameters of proposed model is given in Table 02.

**Table 2:** Proposed Model descriptions.

| Layer | Output | Parameters |
| --- | --- | --- |
| Conv2d (Conv2D) | (None, 222,222,32) | 896 |
| MaxPooling2D | (None, 111,111,32) | 0 |
| Conv2d1 | (None, 109,109,32) | 9248 |
| MaxPooling2D1 | (None, 54,54,32) | 0 |
| dropout (Dropout) | (None, 54,54,32) | 0 |
| Conv2d2 | (None, 52,52,32) | 9248 |
| MaxPooling2D2 | (None, 26,26,32) | 0 |
| dropout1 | (None, 26,26,32) | 0 |
| flatten | (None, 216332) | 0 |
| dense | (None, 128) | 2769024 |
| dropout2 | (None, 128) | 0 |
| Batch normalization | (None, 128) | 512 |
| dense1 | (None, 64) | 8256 |
| dense2 | (None, 3) | 195 |
| activation | (None, 3) | 0 |

Total Parameter: 2,797,379
Trainable Parameter: 2,797,123
Non-Trainable Parameter: 256

## 4 Result and Discussion

### 4.1 Accuracy vs loss plot

Figure 5 shows the average accuracy vs epochs plot while Figure 6 shows average loss vs epochs plot for both training and validation phase for 100 epochs. These curves indicate that the model training progresses without overfitting and hence, the model can be further trained with more data and careful fine tuning. In addition, these curves give accuracy range between 93 to 96%. The detail for accuracies of each class is given in Table 3. It is also pertinent to mention that the accuracy is starting at around 0.65 which is a fairly high value because of a small batch size 32 and high learning rate 0.001. The step per epoch is 370 as the total number of training images are 11844 hence, the model is trained enough in one epoch.

**Fig. 5:**
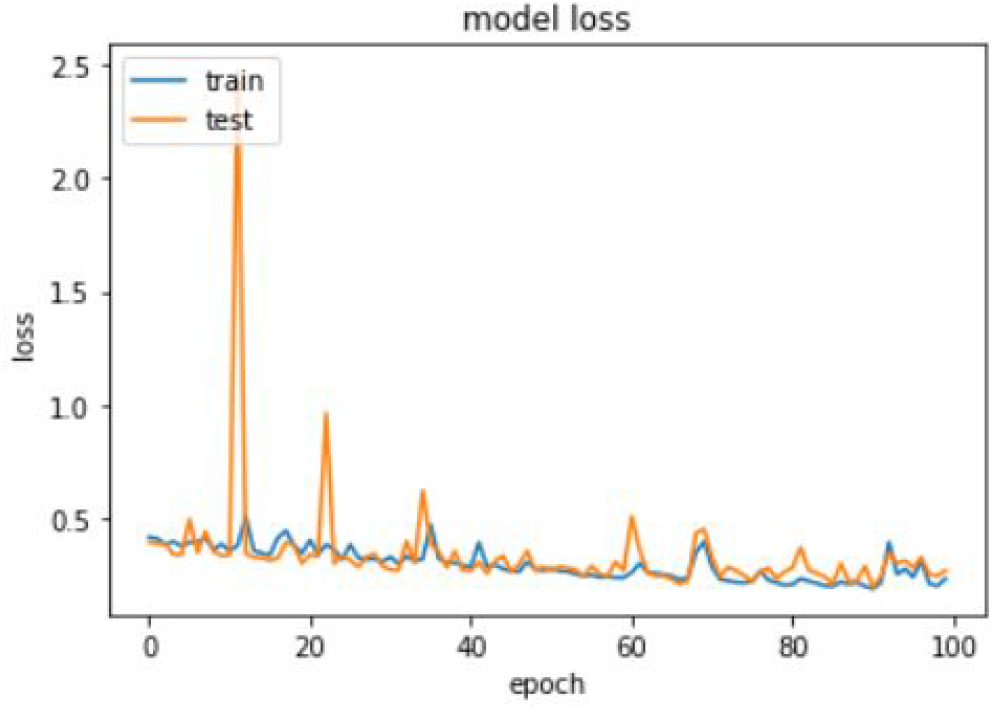
loss vs epochs.

**Fig. 6:**
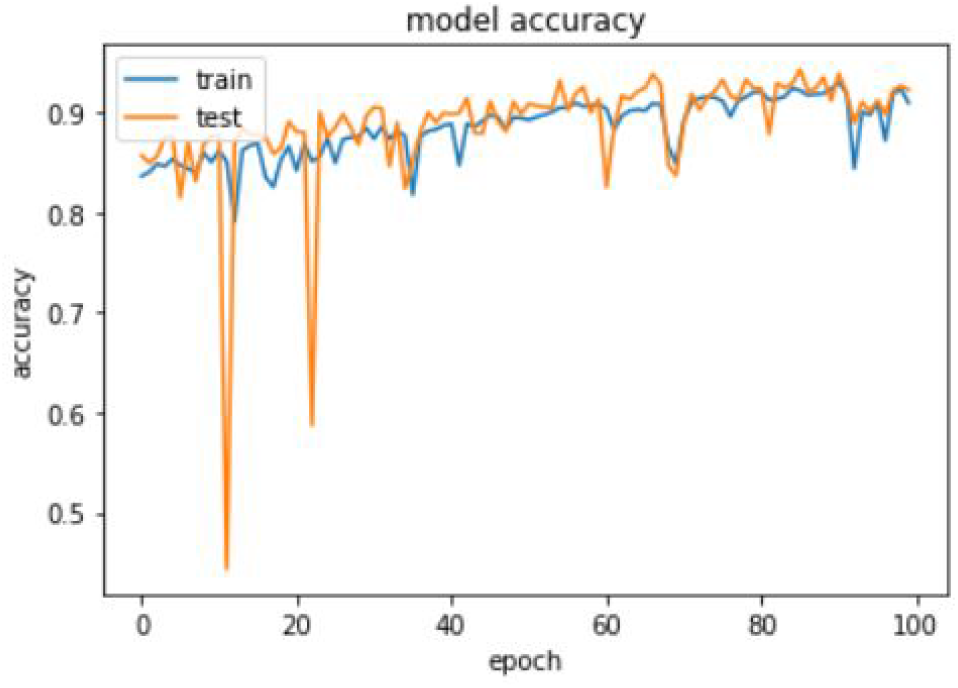
accuracy vs epochs.

**Table 3:**
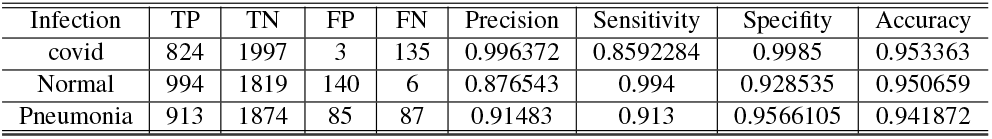
Performance matrices from the confusion matrix of the proposed model

| Infection | TP | TN | FP | FN | Precision | Sensitivity | Specificity | Accuracy |
| --- | --- | --- | --- | --- | --- | --- | --- | --- |
| covid | 824 | 1997 | 3 | 135 | 0.996372 | 0.8592284 | 0.9985 | 0.953363 |
| Normal | 994 | 1819 | 140 | 6 | 0.876543 | 0.994 | 0.928535 | 0.950659 |
| Pneumonia | 913 | 1874 | 85 | 87 | 0.91483 | 0.913 | 0.9566105 | 0.941872 |

### 4.2 Confusion matrix

Confusion matrix (CM) is the backbone of classification performance parameters either in binary classifier or multiclass classifiers. It has four fundamental parameters i.e. True Positive (TP), True Negative (TN), False Positive (FP) and False Negative (FN). CM for the proposed model is given in Figure 7. The predicted label is on horizontal axis while the vertical axis shows true label. Here for COVID-19 class, TP mean the COVID positive cases and they are predicted as positive i.e. 824, TN means the non COVID cases and they are detected as non COVID i.e. 1997, FP is the number of non COVID cases predicted as COVID i.e. 3 and FN is the number of COVID patients predicted as non COVID i.e. 135. The complete detail of confusion matrix for each class is given in Table. 3

**Fig. 7:**
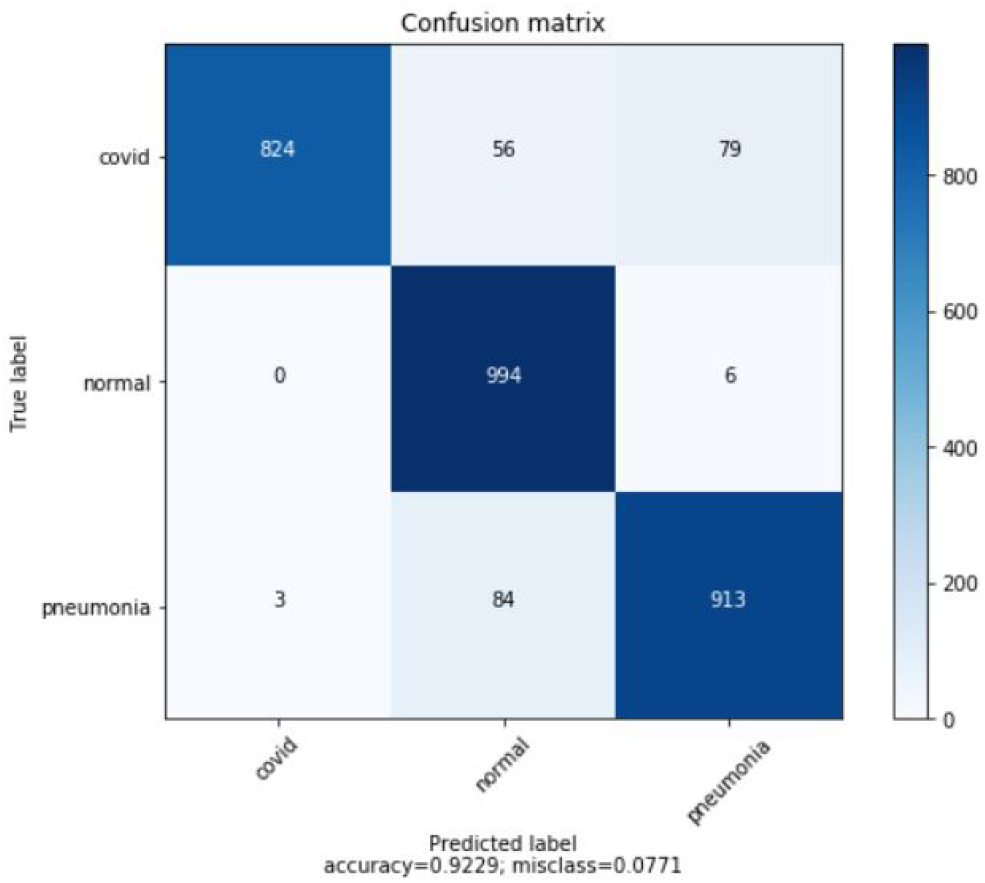
Confusion matrix.

The classification assessment parameters for binary and multiclass classifiers are enlisted in [38] and all of them are based on confusion matrix. The performance assessment of the proposed model is done by using precision, sensitivity, specificity and accuracy. For all classes these parameters are calculated in Table 3 based on the definition of [38]. The proposed model is very prcised for COVID class with precision of 99.96% which mean that the model has excellent performance for COVID cases. Furthermore, the model rarely classifies normal or pneumonia patients as COVID positive. This is also indicated by specificity which is 99.9%. Both of these parameters depend on FP which is 3 for COVID class as shown in the Table 3. Apart from this, the sensitivity of the model is a little bit low i.e. 85.9%, which means that the model classify COVID as normal or as pneumonia because of excessive augmentation used for training purpose. In addition to this, till now the radiological findings i.e. ground glass opacities, consolidation and haziness in chest X-Rays are common between infectious diseases and also the dataset may contain falsely classified COVID cases which cause miss classification. If more data of expert radiologist is available, the sensitivity of the model can be improved for COVID cases.

## 5 Conclusion

Early, economically feasible, accurate detection and classification of COVID-19 by using CXR play an important role to prevent the spread of this global pandemic. Therefore, a simple novel robust deep-learning convolutional neural network (CNN) is proposed which uses a set of 4803 augmented X-rays images of COVID-19, 5000 images of Normal and 5000 images of pneumonia. From these images 80% of each class is used for training the model and remaining 20% images are used for validation purpose. This model classifies COVID-19, Pneumonia and Normal cases with accuracy of 95.33%, 94.18%, 95.06% respectively. Although, the model is simple in architecture, computationally efficient and has promising result but its sensitivity can be improved more if more data of expert radiologist is available.

## Data Availability

All the Datasets are available on the internet

https://www.kaggle.com/alifrahman/covid19-chest-xray-imagedataset

https://www.sirm.org/category/senza-categoria/covid-19/

https://github.com/ieee8023/covid-chestxray-dataset

https://github.com/agchung/Figure1-COVID-chestxray-dataset

https://www.kaggle.com/tawsifurrahman/covid19-radiographydatabase

https://github.com/agchung/Actualmed-COVID-chestxray-dataset

## References

[1] W. H. Organization and W. H. O. World Health Organization Disease outbreak news, “Pneumonia of unknown causeCChina. Emergencies preparedness, response,” 2020.

[2] F. Wu et al., “A new coronavirus associated with human respiratory disease in China,” vol. 579, no. 7798, pp. 265–269, 2020.

[3] www.who.int/emergencies/diseases/novel-coronavirus-2019.

[4] www.who.int/newsroom/detail/07-04-2020-who-lists-two-covid-19-tests-for-emergency-use.

[5] C. Huang et al., “Clinical features of patients infected with 2019 novel coronavirus in Wuhan, China,” vol. 395, no. 10223, pp. 497–506, 2020.

[6] M. Ackermann et al., “Pulmonary vascular endothelialitis, thrombosis, and angiogenesis in Covid-19,” vol. 383, no. 2, pp. 120–128, 2020.

[7] A. Addeo, M. Obeid, and A. J. J. f. i. o. c. Friedlaender, “COVID-19 and lung cancer: risks, mechanisms and treatment interactions,” vol. 8, no. 1, 2020.

[8] Y. Fang et al., “Sensitivity of chest CT for COVID-19: comparison to RT-PCR,” vol. 296, no. 2, pp. E115–E117, 2020.

[9] J. P. Kanne, B. P. Little, J. H. Chung, B. M. Elicker, and L. H. Ketai, “Essentials for radiologists on COVID-19: an updatełradiology scientific expert panel,” ed: Radiological Society of North America, 2020.

[10] X. Xie, Z. Zhong, W. Zhao, C. Zheng, F. Wang, and J. J. R. Liu, “Chest CT for typical coronavirus disease 2019 (COVID-19) pneumonia: relationship to negative RT-PCR testing,” vol. 296, no. 2, pp. E41–E45, 2020.

[11] A. Taghizadieh, A. Ala, F. Rahmani, and A. J. E. Nadi, “Diagnostic accuracy of chest X-ray and ultrasonography in detection of community acquired pneumonia; a brief report,” vol. 3, no. 3, p. 114, 2015.

[12] C. Meng and X. J. I. A. Zhao, “Webcam-based eye movement analysis using CNN,” vol. 5, pp. 19581–19587, 2017.

[13] X. Deng et al., “Joint hand detection and rotation estimation using CNN,” vol. 27, no. 4, pp. 1888–1900, 2017.

[14] A. Krizhevsky, I. Sutskever, and G. E. J. A. i. n. i. p. s. Hinton, “Imagenet classification with deep convolutional neural networks,” vol. 25, pp. 1097–1105, 2012.

[15] M. D. Zeiler and R. Fergus, “Visualizing and understanding convolutional networks,” in European conference on computer vision, 2014, pp. 818–833: Springer.

[16] K. Simonyan and A. J. a. p. a. Zisserman, “Very deep convolutional networks for large-scale image recognition,” 2014.

[17] K. J. R. p. Suzuki and technology, “Overview of deep learning in medical imaging,” vol. 10, no. 3, pp. 257–273, 2017.

[18] G. Litjens et al., “A survey on deep learning in medical image analysis,” vol. 42, pp. 60–88, 2017.

[19] A. S. Lundervold and A. J. Z. f. M. P. Lundervold, “An overview of deep learning in medical imaging focusing on MRI,” vol. 29, no. 2, pp. 102–127, 2019.

[20] I. D. Apostolopoulos, T. A. J. P. Mpesiana, and E. S. i. Medicine, “Covid-19: automatic detection from x-ray images utilizing transfer learning with convolutional neural networks,” vol. 43, no. 2, pp. 635–640, 2020.

[21] A. Narin, C. Kaya, and Z. J. a. p. a. Pamuk, “Automatic detection of coronavirus disease (covid-19) using x-ray images and deep convolutional neural networks,” 2020.

[22] J. Zhang, Y. Xie, Y. Li, C. Shen, and Y. J. a. p. a. Xia, “Covid-19 screening on chest x-ray images using deep learning based anomaly detection,” 2020.

[23] P. K. Sethy, S. K. Behera, P. K. Ratha, and P. Biswas, “Detection of coronavirus disease (COVID-19) based on deep features and support vector machine,” 2020.

[24] L. O. Hall, R. Paul, D. B. Goldgof, and G. M. J. a. p. a. Goldgof, “Finding covid-19 from chest x-rays using deep learning on a small dataset,” 2020.

[25] L. Li et al., “Artificial intelligence distinguishes COVID-19 from community acquired pneumonia on chest CT,” 2020.

[26] Wang L., Wong A.. COVID-Net: A tailored deep convolutional neural network design for detection of COVID-19 cases from chest radiography images. arXiv preprint 2003098712020

[27] E. E.-D. Hemdan, M. A. Shouman, and M. E. J. a. p. a. Karar, “Covidx-net: A framework of deep learning classifiers to diagnose covid-19 in x-ray images,” 2020.

[28] Y. Song et al., “Deep learning enables accurate diagnosis of novel coronavirus (COVID-19) with CT images,” 2020.

[29] M. Barstugan, U. Ozkaya, and S. J. a. p. a. Ozturk, “Coronavirus (covid-19) classification using ct images by machine learning methods,” 2020.

[30] M. M. Ahsan, T. E Alam, T. Trafalis, and P. J. S. Huebner, “Deep MLP-CNN model using mixed-data to distinguish between COVID-19 and Non-COVID-19 patients,” vol. 12, no. 9, p. 1526, 2020.

[31] www.kaggle.com/alifrahman/covid19-chest-xray-image-dataset.

[32] www.kaggle.com/c/rsna-pneumonia-detection-challenge.

[33] www.sirm.org/category/senza-categoria/covid-19/.

[34] github.com/ieee8023/covid-chestxray-dataset.

[35] github.com/agchung/Figure1-COVID-chestxray-dataset.

[36] www.kaggle.com/tawsifurrahman/covid19-radiography-database.

[37] github.com/agchung/Actualmed-COVID-chestxray-dataset.

